# Endovascular thrombectomy with and without preceding thrombolysis in posterior circulation stroke – insights from STAR

**DOI:** 10.1101/2024.02.22.24303230

**Authors:** Ev-Christin Heide, Sami Al Kasab, Ali Alawieh, Adam Arthur, Waleed Brinjikji, Shakeel Chowdhry, Roberto Crosa, Hugo Cuellar, Reade De Leacy, Travis Dumont, Marielle Ernst, Mohamad Ezzeldin, Isabel Fragata, Brian Howard, Pascal Jabbour, Peter Kan, Joon-Tae Kim, Michael R. Levitt, Justin Mascitelli, Charles Matouk, Mark Moss, Pedro Navia, Joshua Osbun, Min S. Park, Adam Polifka, Marios-Nikos Psychogios, Ansaar Rai, Daniele G. Romano, Amir Shaban, Robert M. Starke, Omar Tanweer, Richard Williamson, Stacey Q. Wolfe, Shinichi Yoshimura, Alejandro M. Spiotta, Ilko L. Maier, STAR Investigators

## Abstract

**Background:** Multiple randomized trials could not establish the non-inferiority of endovascular thrombectomy (EVT) alone without preceding intravenous thrombolysis (IVT) or superiority of IVT followed by EVT in anterior circulation large vessel occlusion (LVO) stroke. The role of prior IVT in posterior circulation LVO remains controversial.

**Methods:** In this multicenter, retrospective study, stroke patients with LVO in the posterior circulation who received EVT alone or with IVT were selected from the stroke thrombectomy and aneurysm registry (STAR) between 2013 to 2022. Effects of IVT followed by thrombectomy on favorable functional outcome (defined as modified Rankin scale ≤ 3 at 90 days) and safety were investigated using multivariable logistic and linear regression models.

**Results:** Of the 588 included patients, 67 % (n = 394) were treated with EVT alone and 29% (n = 170) with EVT after IVT and 4% (n = 24) have missing values on this variable. Controlling for multiple confounding factors, IVT was not associated with a higher likelihood of favorable functional outcome at 90 days (odds ratio 1.04, 95 % CI 0.52-2.09, p = 0.901). Thrombectomy alone did not show any safety advantages compared with those receiving IVT.

**Conclusions:** Similar functional outcomes and complication rates were seen in patients with posterior circulation LVO treated with EVT alone vs EVT after IVT. Further prospective studies are required to determine the utility of IVT in posterior circulation stroke, especially in patients being directly admitted to thrombectomy centers.

## Introduction

Endovascular thrombectomy (EVT) in combination with best medical treatment has been shown to improve functional outcomes and reduce mortality rates compared with medical treatment alone in acute basilar artery (BA) occlusion stroke.^1,2^ However, the effect of EVT with prior intravenous thrombolysis (IVT) versus EVT alone in stroke from large vessel occlusion (LVO) of the posterior circulation (PC) remains unclear. IVT prior to EVT may improve recanalization rates and functional outcomes, but may also increase the risk of clot fragmentation with distal embolization, delay interventional treatment, add costs, and increase the risk of intracranial hemorrhage.^3,4^

The non-inferiority of IVT followed by thrombectomy for anterior circulation LVO has been evaluated in six multicenter, randomized controlled clinical trials: Two trials showed non-inferiority of EVT alone^5,6^, while four trials failed to demonstrate non-inferiority.^7–10^ A recent meta-analysis of these six studies could neither establish superiority of IVT plus EVT nor non-inferiority of EVT alone in patients presenting directly at endovascular treatment centers with anterior circulation LVO. Differences in functional outcomes between treatment groups were small and not significant.^11^ On the basis of this meta-analysis, the loss of a clinically relevant effect when omitting IVT before EVT in patients presenting directly at centers capable of endovascular treatment cannot be excluded. However, the benefit of IVT in this setting seems to be small. In contrast, in patients with secondary transfers to thrombectomy centers (‘drip-and-ship scenari’) randomized trials on the benefit of IVT before EVT are missing likely due to a real equipoise.

To date, no randomized trials have investigated the role of IVT followed by thrombectomy in PC-LVO. In a retrospective study of 322 patients with acute BA occlusion, functional outcomes were similar in patients treated with IVT versus EVT alone.^12^ However, in patients with underlying large-artery atherosclerosis, a significant beneficial treatment effect of IVT followed by thrombectomy was observed. Conversely, a meta-analysis of 1096 patients demonstrated that IVT followed by thrombectomy was associated with lower mortality rates at 90 days compared with EVT alone in patients with acute BA occlusion, while no differences have been observed concerning functional outcomes or rates of intracranial hemorrhages.^13^

The aim of this study is to investigate the effect of IVT prior to EVT in patients with PC-LVO compared to EVT alone using a large, international ‘real world’ stroke database. Given the retrospective nature of this study, the results are purely observational and to be interpreted as hypotheses-generating to encourage further prospective studies.

## Methods

### Patient cohort

Patients with acute PC-LVO who underwent EVT with or without systemic thrombolysis between 2013 and 2022 were identified using a multicenter, international database. The Stroke Thrombectomy and Aneurysm Registry (STAR) includes a diverse cohort of individuals who have undergone EVT, reflecting real-world clinical practice.^14^ STAR comprises 85 stroke centers worldwide, in which EVT is performed on a regular basis following highest standards. Data from this registry contains preinterventional, intrainterventional, and postinterventional, prospectively documented information of treatment periods, prestroke functional status, medical history, interventional strategy, and used materials as well as functional outcome using the National Institutes of Health Stroke Scale (NIHSS; baseline and at discharge) and modified Rankin scale (mRS; at discharge and at 90 days).

For this study, we selected patients with following inclusion criteria: (1) BA occlusion with or without extension into one or both posterior cerebral arteries, (2) no acute intracranial stenting performed, (3) no treatment with intraarterial tissue-type plasminogen activator (IA-tPA). All patients with other occlusions than BA or basilar-posterior artery occlusion in the primary/first vessels for which thrombectomy was attempted, or patients with intracranial stenosis that needed intervention (Percutaneous transluminal angioplasty, stenting, or acute surgery) and intra-arterial (IA) thrombolysis or spontaneous reperfusion without EVT were excluded.

The primary outcome was favorable functional outcome defined as modified Rankin scale [mRS] ≤ 3 at 90 days. Secondary outcomes included mortality by 90 days, mRS ≤ 3 at discharge, successful reperfusion (defined as good final modified Thrombolysis in Cerebral Infarction score [mTICI ≥ 2b]), number of recanalization attempts, NIHSS at discharge, change in NIHSS between presentation and discharge and complications.

The data that support the findings of this study are available from the corresponding author upon reasonable request. This study was approved by institutional review boards at each study center, and there was no need for written informed consent given the retrospective design of the study. The study follows the “Strengthening the Reporting of Observational Studies in Epidemiology” statement.^15^ The data that support the findings of this study are available from the corresponding author on reasonable request.

### Statistical analysis

Statistical analyses were performed using Stata 17.0. (StataCorp., College Station, Texas). Characteristics of all patients are shown as mean ± standard deviation (SD), if normally distributed, and as median with interquartile range for non-parametric data. P-values < 0.05 were considered statistically significant. The analysis of interest is a comparison of outcomes by cohort (EVT alone vs IVT + EVT). Multiple imputation was used to address missingness prior to analysis. Multiple imputation was performed using the chained equations approach.^16^ Forty imputations were used for this analysis. The N for all analyses is 588. For the analysis model, logistic regression was used to estimate analysis models for binary outcomes (mRS at 90 days, mortality by 90 days, mRS ≤ 3 at discharge, successful reperfusion, and complications). Final mTICI and mRS at 90 days were analyzed using ordered logistic regression. Based on the nature of the variables and their distributions negative binomial models were used for number of total recanalization attempts, length of stay and time to successful reperfusion. Results of all analyses that used logistic, ordered logistic, or negative binomial regression are reported both as average marginal effects, and/or exponentiated coefficients (odds ratios [OR] or incidence-rate ratios). Linear models were used for outcomes NIHSS at discharge and 90 days, and change in NIHSS between admission and discharge. Results of analyses that used linear models are reported as linear regression coefficients. Variables that are associated with both exposure (IVT) and outcome, but do not lie on the causal path from exposure to outcome, were defined as confounders. For all the analyses, the following confounders were included: age, sex, race, history of prior stroke, history of hypertension, history of diabetes, history of atrial fibrillation, history of hyperlipidemia, former smoker, current smoker, history of congestive heart failure. The confounders are displayed in the Supplemental Material. Due to the observational nature of this study, they cannot be interpreted as describing causal effects.

## Results

### Characteristics of patients and time metrics

588 patients with PC-LVO treated with EVT were identified from the STAR database. Of these patients, 67 % (n = 394) were treated with EVT alone and 29% (n = 170) with IVT followed by EVT and in 4% (n = 24) IVT status was unknown. The characteristics of the patients at baseline are presented in Table 1. The proportion of missing values for variables in this study ranged from 0 % to 65 % (Supplemental Table 1). There were no differences in age, sex, vascular risk factors, history of prior stroke, type of occlusion (basilar occlusion alone, or basilar occlusion with additional posterior cerebral artery occlusion) between groups. The median NIHSS on admission was 17 (IQR 8-26) in the EVT alone group and 19 (IQR 11-26) in the IVT group. In the EVT alone group, the median time from puncture to recanalization was 39 (IQR 22-68) minutes (min) compared to 36 (IQR 26-60) min in the IVT group. The median time from symptom onset or last known well to puncture differed by 245 min between the groups (EVT alone: 503 [288-850] min and IVT: 258 [180-340] min).

**Table 1:**
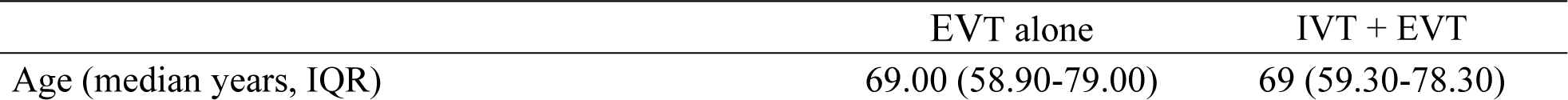
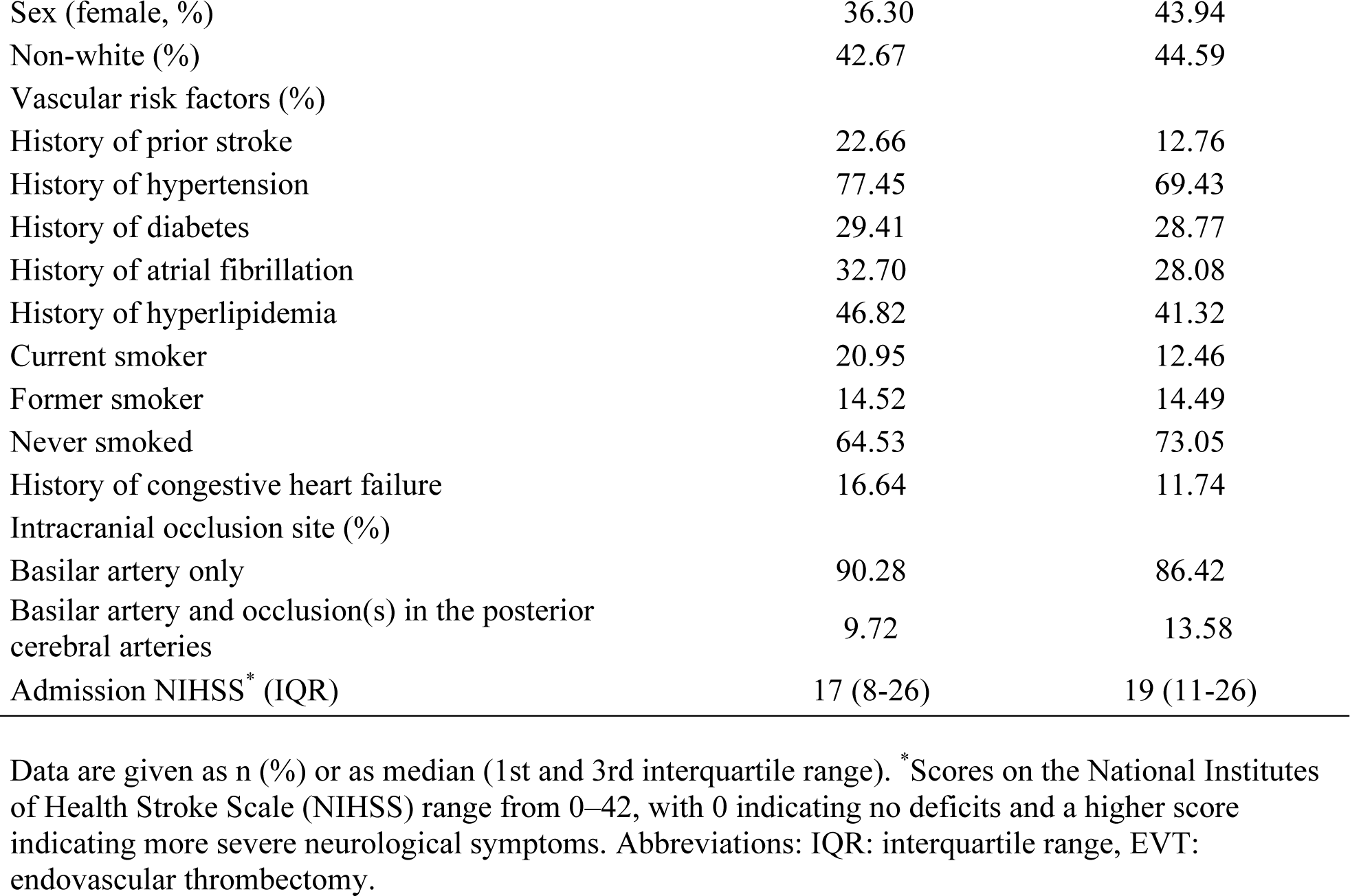
Baseline characteristics of patient with posterior circulation large vessel occlusion with- and without intravenous thrombolysis (IVT) prior to endovascular thrombectomy (EVT).

|  | EVT alone | IVT + EVT |
| --- | --- | --- |
| Age (median years, IQR) | 69.00 (58.90-79.00) | 69 (59.30-78.30) |
| Sex (female, %) | 36.30 | 43.94 |
| Non-white (%) | 42.67 | 44.59 |
| Vascular risk factors (%) |  |  |
| History of prior stroke | 22.66 | 12.76 |
| History of hypertension | 77.45 | 69.43 |
| History of diabetes | 29.41 | 28.77 |
| History of atrial fibrillation | 32.70 | 28.08 |
| History of hyperlipidemia | 46.82 | 41.32 |
| Current smoker | 20.95 | 12.46 |
| Former smoker | 14.52 | 14.49 |
| Never smoked | 64.53 | 73.05 |
| History of congestive heart failure | 16.64 | 11.74 |
| Intracranial occlusion site (%) |  |  |
| Basilar artery only | 90.28 | 86.42 |
| Basilar artery and occlusion(s) in the posterior cerebral arteries | 9.72 | 13.58 |
| Admission NIHSS* (IQR) | 17 (8-26) | 19 (11-26) |

Data are given as n (%) or as median (1st and 3rd interquartile range). \* Scores on the National Institutes of Health Stroke Scale (NIHSS) range from 0–42, with 0 indicating no deficits and a higher score indicating more severe neurological symptoms. Abbreviations: IQR: interquartile range, EVT: endovascular thrombectomy.

### Primary outcomes

There was no difference in rates of favorable functional outcome between groups at 90 days (OR 1.02, 95% confidence interval [CI] 0.57-1.82; Figure 1 and Table 2). The primary outcome was reached in 45.66 % of patients in the EVT alone group and in 46.50 % of the patients in the IVT group.

**Figure 1:**
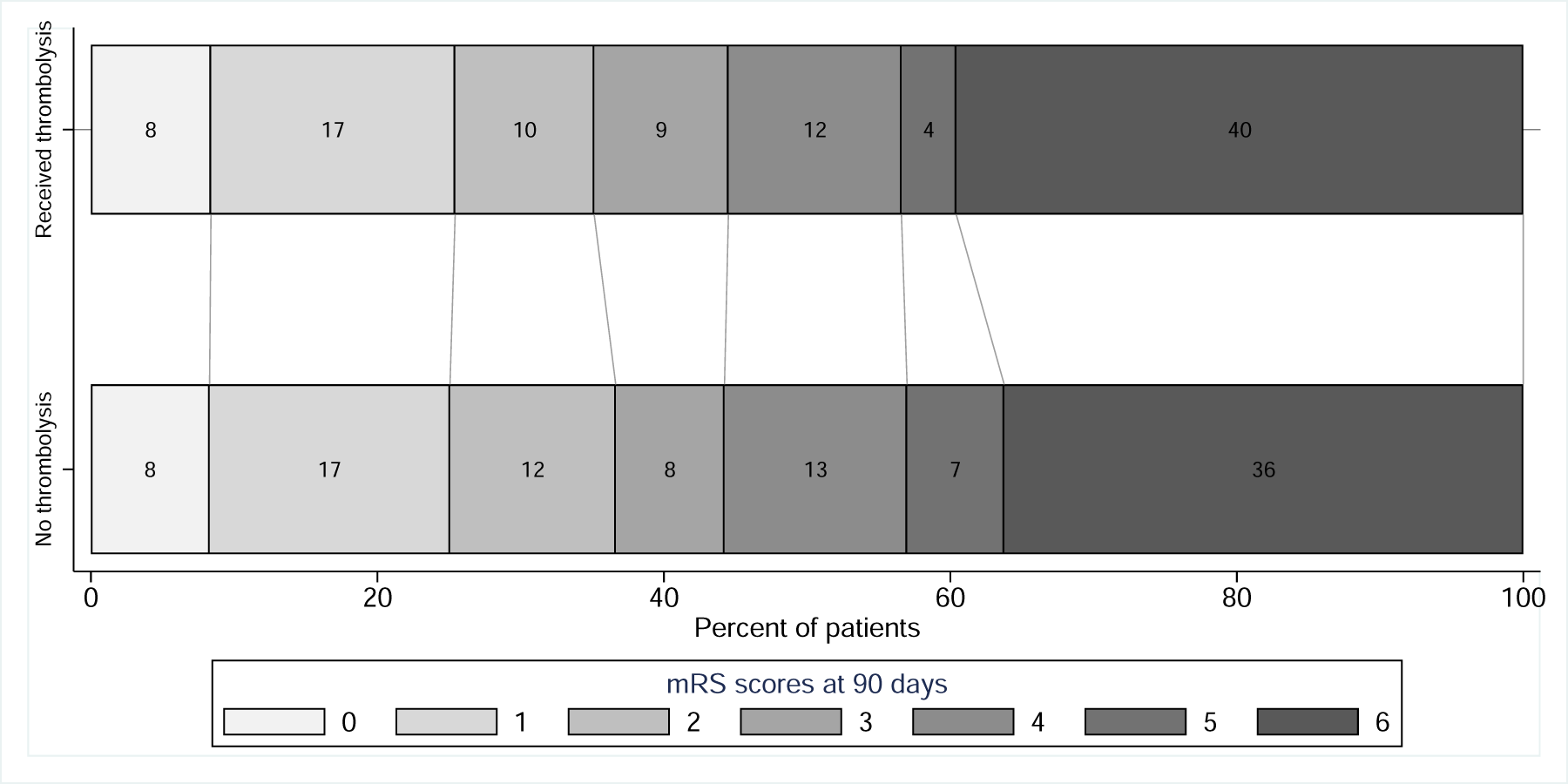
Shift plot of modified Rankin scale (mRS) at 90 days. The primary endpoint (mRS ≤3) was reached in 45.66 % and 46.50 % of the patients in the endovascular thrombectomy alone group and the intravenous thrombolysis group, respectively (odds ratio 1.02, 95% confidence interval 0.57-1.82, p = 0.957).

**Table 2:** Multivariable logistic regression for favorable functional outcome (mRS^†^ at 90 days) in patients with posterior circulation large vessel occlusion with and without intravenous thrombolysis prior to thrombectomy.

|  | OR (95 % CI) | P-value | AME (95 % CI) | P-value |
| --- | --- | --- | --- | --- |
| Received intravenous thrombolysis | 1.02 (0.57-1.82) | 0.957 | < 0.01 (-0.13-0.13) | 0.951 |

### Secondary and safety outcomes

Secondary clinical and technical efficacy outcomes are shown in Table 3. There was no significant difference between the groups in successful reperfusion, 90-day mortality, mRS ≤ 3 at discharge, NIHSS at discharge, or successful reperfusion rate after completion of thrombectomy (≥ mTICI2b). The median number of recanalization attempts was significantly higher in the EVT alone group (2 [1-3] attempts) compared to the IVT group (1 [1-2] attempts; incidence-rate ratio 0.84, 95 % CI 0.72-0.98, p = 0.023). There was a trend towards a greater reduction in NIHSS between admission and discharge in the IVT group (EVT alone: reduction of 5 points [0-12]; IVT: reduction of 7 points [1-17] points, linear regression coefficient −2.805, 95 % CI −6.05-0.44, p = 0.089).

**Table 3:** Regression of secondary outcome parameters in patients with posterior circulation large vessel occlusion with- and without intravenous thrombolysis prior to thrombectomy.

**Ordered logistic regression**
|  | OR (95 % CI) | P-value | AME (95 % CI) | P-value |
| --- | --- | --- | --- | --- |
| Mortality by 90 days | 1.18 (0.72-1.92) | 0.516 | 0.04 (-0.07- 0.15) | 0.518 |
| mRS<sup>†</sup> at discharge $\leq 3$ | 1.12 (0.73-1.71) | 0.607 | 0.03 (-0.07-0.12) | 0.608 |
| mTICI<sup>o‡</sup> $\geq 2b$ | 0.82 (0.49-1.35) | 0.435 | -0.03 (-0.10-0.04) | 0.444 |
| mRS<sup>†</sup> at 90 days | 1.13 (0.72-1.79) | 0.593 |  |  |
| 0 |  |  | -0.01 (-0.04-0.02) | 0.604 |
| 1 |  |  | -0.01 (-0.06-0.03) | 0.602 |
| 2 |  |  | -0.01 (-0.02-0.01) | 0.583 |
| 3 |  |  | < 0.01 (-0.01-0.01) | 0.594 |
| 4 |  |  | > -0.01 (> -0.01-< 0.01) | 0.952 |
| 5 |  |  | < 0.01 (> -0.01-0.01) | 0.604 |
| 6 |  |  | 0.03 (-0.07-0.13) | 0.596 |

**Negative binomial regression**
|  | Incidence-rate ratio<br>(95 % CI) | P-value | AME (95 % CI) | P-value |
| --- | --- | --- | --- | --- |
| Recanalization attempts | 0.84 (0.72-0.98) | 0.023* | -0.36 (-0.65- -0.06) | 0.019* |

**Linear regression**
|  | Linear regression coefficient<br>(95 % CI) | P-value |
| --- | --- | --- |
| Median NIHSS<sup>§</sup> at discharge | -1.20 (-4.11-1.71) | 0.417 |
| Change in NIHSS<sup>§</sup> (discharge – admission) | -2.80 (-6.05-0.44) | 0.089 |

\*p < 0.05. <sup>†</sup>Score on the modified Rankin scale (mRS) range from 0 (no symptoms) to 6 (death).
<sup>‡</sup>Grades on the modified Thrombolysis in Cerebral Infarction Scale (mTICI) range from 0 (no reperfusion) to 3 (complete reperfusion). <sup>§</sup>Scores on the National Institutes of Health Stroke Scale (NIHSS) range from 0–42, with 0 indicating no deficits and a higher score indicating more severe neurological symptoms. Abbreviations: CI: confidence interval, min: minute(s).

Complications are shown in Table 4. The risks for symptomatic intracranial hemorrhage (sICH) were inconclusive. Whereas the average marginal effect (AME) was significantly different with higher rates in the EVT alone group (5.79 %) compared to the IVT group (2.78 %; AME −0.04, 95 % CI −0.07 - <0.01, p = 0.043), the OR was without significant differences between the two groups (OR 0.37, 95 % CI 0.12-1.16, p = 0.088). There was no significant difference in overall complications, hemorrhage or distal embolization between the two groups.

**Table 4:** Logistic regression of safety outcome parameters in patients with posterior circulation large vessel occlusion with- and without intravenous thrombolysis prior to thrombectomy.

|  | OR (95 % CI) | P-value | AME (95 % CI) | P-value |
| --- | --- | --- | --- | --- |
| Any complication | 0.76 (0.38-1.49) | 0.419 | -0.02 (-0.08-0.03) | 0.403 |
| Distal embolization | 1.87 (0.88-3.97) | 0.105 | 0.06 (-0.02-0.13) | 0.125 |
| Hemorrhage | 0.92 (0.56-1.51) | 0.750 | -0.01 (-0.08-0.06) | 0.748 |
| Symptomatic intracerebral hemorrhage | 0.37 (0.12-1.16) | 0.088 | -0.04 (-0.07- <0.01) | 0.043* |

## Discussion

This large, international, multicenter study compared EVT alone to IVT followed by thrombectomy in a cohort of patients with posterior circulation LVO. There was no significant difference in the 90-day mRS between the two groups.

There are no randomized controlled studies on the effect of IVT followed by EVT in posterior circulation LVO; three multicenter studies, one cohort study and multiple meta-analyses have reported equivocal results. To our knowledge, our study is the largest (n = 588). In accordance with our study, Siow et al. found similar overall functional outcomes and mortality in BA occlusion patients treated with EVT alone or IVT followed by EVT.^12^ However, in patients with underlying BA atherosclerosis, IVT conferred significant benefit. One limitation of our study is the lack of stroke etiology, so this subgroup could not be analyzed in our cohort. Similarly, Nappini et al. studied 464 patients from a national prospective registry and found no difference in successful reperfusion (mTICI ≥ 2b), functional outcomes and mortality between EVT alone and IVT followed by EVT.^17^ In contrast, Nie et al. found improved functional outcomes without differences in reperfusion or mortality in patients with IVT followed by EVT compared to EVT alone.^18^ In a meta-analysis of these three existing but conflicting studies and one additional retrospective institutional study, IVT followed by thrombectomy was associated with significantly higher odds of superior functional outcomes at 90 days and lower mortality rates than EVT alone.^19^ In our study, we could not find a significant difference in the functional outcome at 90 days between the two treatment groups. Thus, our study questions the benefit of IVT followed by thrombectomy. All these retrospective studies did not differentiate between mothership and drip-and-ship-patients, which might explain the varying results. Nevertheless, in our cohort, a potential beneficial effect of IVT followed by thrombectomy did not seem to be strong. Despite an earlier treatment and no underlying contraindications for IVT we could not find a significant difference in the functional outcome. In a meta-analysis of six randomized clinically trials, the non-inferiority of IVT followed by thrombectomy for anterior circulation LVO was studied.^11^ The loss of a clinically relevant effect when omitting IVT before EVT in patients presenting directly at centers capable of endovascular treatment cannot be excluded. However, the benefit of IVT followed by thrombectomy in this setting also seems to be small.

The trend for a greater reduction in NIHSS between admission and discharge in the IVT group was not significant and might be of less importance in the light of similar long-term outcomes. In addition, the change in NIHSS could be explained by the significantly shorter interval between stroke symptom onset and puncture time in the IVT group and thus is most likely a selection bias. Previous studies reported a significantly longer interval between stroke symptom onset and puncture time in patients treated with EVT alone compared to IVT followed by thrombectomy as well (Siow et al.: 330 min vs. 240 min [mean] and Kohli et al.: 711 min vs. 313 min [median]).^13,19^ Multiple regression models cannot fully account for the longer time interval between stroke onset and puncture in the EVT alone group, which might have influenced the outcomes and is a limitation of this study. Interestingly, Nie et al.^18^ reported an improved functional outcome in the IVT group despite the highest median onset to puncture time of 510 min in the IVT group (IQR 378-763). Concerning the onset to puncture time of the IVT group in our study (median and IQR: 258 min [180-340 min]), it was similar in comparison with Siow et al. (mean and SD: 240 ± 60 min) and Nappini et al. (median and IQR: 280 min [208-370 min]).^12,17^ The time from puncture to recanalization was smaller in our cohort compared to Siow et al. (mean ± SD: EVT group 60 min ± 30 min vs. IVT group 60 min ± 30 min) and to Nie et al. (median [IQR]: EVT group 122 min [85-175 min] vs. IVT group 122 min [60-185 min]). Most importantly, the puncture to recanalization time was not significantly different between the two treatment groups in our study and all referenced studies.^12,17,18^

Our results were inconclusive concerning the rate of sICH between the groups. Whereas AME was significantly different with slightly higher rates of sICH in the EVT group, the OR was without significant difference between the treatment groups. One could speculate that a higher rate of sICH could be explained by the significantly higher number of recanalization attempts in the sole EVT group in our cohort, raising the risk of vessel wall injury and rupture. In any case, IVT followed by thrombectomy was not associated with a higher sICH rate. In line with this, sICH rates were similar among the treatment groups in the above-mentioned studies.^12,17,18^

It is assumed that IVT leads to an improved and earlier recanalization due to clot softening and to an improved microvascular reperfusion due to dissolving of distal emboli, that are inaccessible to thrombectomy devices.^4^ We found a significantly reduced number of recanalization attempts in the IVT group in our cohort with an effect size of one attempt. This is in contrast to reported drawbacks of IVT including distal embolization, sICH, and delayed EVT.^4,20^ However, a similar safety outcome would not justify a treatment without differences in mortality or long-term outcome. The lower number of recanalization attempts could also be attributed to other factors like stroke etiology (e.g. Intracranial Atherosclerotic Disease or cardiac embolism) or the shorter time from onset to recanalization in the IVT group.^21^

Our study has several limitations. First, the retrospective design introduces selection bias, and radiographic and clinical results were self-adjudicated. Multivariable analyses cannot adjust completely for systematic differences between treatment groups without a randomized trial design. In addition, other comorbidities and information on contraindications for IVT such as anticoagulants and blood pressure levels or detailed imaging data were not listed in the registry and thus could not be accounted for. However, comorbidities often represent a contraindication to IVT and might have biased the results. Furthermore, there was no information on stroke etiology in our study, precluding subgroup analysis of patients with underlying atherosclerosis. Second, the time from stroke onset to puncture was higher in the EVT alone group which could have reduced the reversibility of the clinical symptoms. However, we chose a multivariable analysis which accounted for this variable. Although the inclusion criteria for EVT are likely to be dependent on the individual centers, it is very likely that in late time windows the dawn and defuse criteria have been used, also identifying patients likely to experience favorable outcomes even when presenting in the late time window. The point of individual inclusion and exclusion criteria based on the SOPs of every individual center on the one hand can be a limitation, but on the other also represents current practice in multiple centers worldwide and therefore real word data. Third, there was no information if the patients received thrombolytic treatment in mothership, drip-and-ship, or ship-only paradigms. Most probably, patients from all paradigms were included. However, patients receiving the mothership protocol are probably the most interesting cohort regarding this question. Therefore, there is a need for prospective randomized controlled clinical trials investigating the effect of IVT in patients with posterior circulation LVO receiving mothership protocol only. Finally, there was a certain missingness of values. Multiple imputation was used to address missingness prior to analysis. Nevertheless, this might have influenced the results.

In conclusion, our large multicenter study of EVT alone compared to IVT followed by thrombectomy in patients with posterior circulation LVO showed similar long-term functional outcomes and complication rates. Given the retrospective study design, we cannot rule out possible beneficial effects of IVT in a subgroup of patients (drip-and-ship-patients). However, our study generally questions the benefit of IVT in patients before EVT and underlines the need of randomized clinical trials to answer this question. Maybe investigating superiority of thrombolysis might be the better trial design compared to a non-inferiority design.

## Acknowledgments

We thank R. A. Medeiros (engaged by STAR) for her statistical support.

## Data availability

The data that support the findings of this study are available from the corresponding author upon reasonable request.

## Author Contributions

Conception and design of the study: IM, ECH; Analysis of data: ECH, IM; Drafting of the manuscript: ECH and IM; Drafting of the figures: ECH, IM; all authors critically revised and approved the final version of the manuscript.

## Supplemental Material

Supplemental Methods: Tables S1-S13

## Nonstandard abbreviations and acronyms

AME: Average marginal effect
BA: Basilar artery
CI: Confidence interval
IA: Intra-arterial
IA-tPA: Intraarterial tissue-type plasminogen activator
IQR: Interquartile range
IVT: Intravenous thrombolysis
LVO: Large vessel occlusion
mRS: Modified Rankin scale
EVT: Endovascular thrombectomy
NIHSS: National institutes of health stroke scale
OR: Odds ratio
PC: Posterior circulation
SD: Standard deviation
sICH: Symptomatic intracranial hemorrhage
STAR: The stroke thrombectomy and aneurysm registry
mTICI: Modified Thrombolysis in Cerebral Infarction

## References

1. Zi W, Qiu Z, Wu D, Li F, Liu H, Liu W, Huang W, Shi Z, Bai Y, Liu Z, et al. Assessment of Endovascular Treatment for Acute Basilar Artery Occlusion via a Nationwide Prospective Registry. JAMA Neurol. 2020;77:561–573.

2. Zhao Y, Zhao W, Guo Y, Li Y. Endovascular thrombectomy versus standard medical treatment for stroke patients with acute basilar artery occlusion: a systematic review and meta-analysis. J Neurointerv Surg. 2022;14:1173–1179.

3. Desilles JP, Loyau S, Syvannarath V, Gonzalez-Valcarcel J, Cantier M, Louedec L, Lapergue B, Amarenco P, Ajzenberg N, Jandrot-Perrus M, et al. Alteplase Reduces Downstream Microvascular Thrombosis and Improves the Benefit of Large Artery Recanalization in Stroke. Stroke. 2015;46(11):3241–8.

4. Fischer U, Kaesmacher J, Mendes Pereira V, Chapot R, Siddiqui AH, Froehler MT, Cognard C, Furlan AJ, Saver JL, Gralla J. Direct Mechanical Thrombectomy Versus Combined Intravenous and Mechanical Thrombectomy in Large-Artery Anterior Circulation Stroke: A Topical Review. Stroke. 2017;48:2912–2918.

5. Yang P, Zhang Y, Zhang L, Zhang Y, Treurniet KM, Chen W, Peng Y, Han H, Wang J, Wang S, et al. Endovascular Thrombectomy with or without Intravenous Alteplase in Acute Stroke. N Engl J Med. 2020;382:1981–1993.

6. Zi W, Qiu Z, Li F, Sang H, Wu D, Luo W, Liu S, Yuan J, Song J, Shi Z, et al. Effect of Endovascular Treatment Alone vs Intravenous Alteplase Plus Endovascular Treatment on Functional Independence in Patients With Acute Ischemic Stroke: The DEVT Randomized Clinical Trial. JAMA. 2021;325:234–243.

7. Suzuki K, Matsumaru Y, Takeuchi M, Morimoto M, Kanazawa R, Takayama Y, Kamiya Y, Shigeta K, Okubo S, Hayakawa M, et al. Effect of Mechanical Thrombectomy Without vs With Intravenous Thrombolysis on Functional Outcome Among Patients With Acute Ischemic Stroke: The SKIP Randomized Clinical Trial. JAMA. 2021;325:244–253.

8. LeCouffe NE, Kappelhof M, Treurniet KM, Rinkel LA, Bruggeman AE, Berkhemer OA, Wolff L, van Voorst H, Tolhuisen ML, Dippel DWJ, et al. A Randomized Trial of Intravenous Alteplase before Endovascular Treatment for Stroke. N Engl J Med. 2021;385:1833–1844.

9. Fischer U, Kaesmacher J, Strbian D, Eker O, Cognard C, Plattner PS, Bütikofer L, Mordasini P, Deppeler S, Pereira VM, et al. Thrombectomy alone versus intravenous alteplase plus thrombectomy in patients with stroke: an open-label, blinded-outcome, randomised non-inferiority trial. Lancet. 2022;400:104–115.

10. Mitchell PJ, Yan B, Churilov L, Dowling RJ, Bush S, Nguyen T, Campbell BCV, Donnan GA, Miao Z, Davis SM. DIRECT-SAFE: A Randomized Controlled Trial of DIRECT Endovascular Clot Retrieval versus Standard Bridging Therapy. J Stroke. 2022;24:57–64.

11. Majoie CB, Cavalcante F, Gralla J, Yang P, Kaesmacher J, Treurniet KM, Kappelhof M, Yan B, Suzuki K, Zhang Y, et al. Value of intravenous thrombolysis in endovascular treatment for large-vessel anterior circulation stroke: individual participant data meta-analysis of six randomised trials. Lancet. 2023;402(10406):965–974.

12. Siow I, Tan BYQ, Lee KS, Ong N, Toh E, Gopinathan A, Yang C, Bhogal P, Lam E, Spooner O, et al. Bridging Thrombolysis versus Direct Mechanical Thrombectomy in Stroke Due to Basilar Artery Occlusion. J Stroke. 2022;24:128–137.

13. Lee KS, Siow I, Zhang JJ, Syn NL, Gillespie CS, Yuen LZ, Anil G, Yang C, Chan BP, Sharma VK, et al. Bridging thrombolysis improves survival rates at 90 days compared with direct mechanical thrombectomy alone in acute ischemic stroke due to basilar artery occlusion: a systematic review and meta-analysis of 1096 patients. J Neurointerv Surg. 2022.

14. Alawieh AM, Spiotta AM. The Stroke Thrombectomy and Aneurysm Registry: Inception, Present, and Future. World Neurosurg. 2020;138:562–564.

15. Elm E von, Altman DG, Egger M, Pocock SJ, Gøtzsche PC, Vandenbroucke JP. The Strengthening the Reporting of Observational Studies in Epidemiology (STROBE) statement: guidelines for reporting observational studies. Lancet. 2007;370:1453–1457

16. Little RJA, Rubin DB. Statistical analysis with missing data. 2019. 3rd ed. Hoboken, NJ: Wiley.

17. Nappini S, Arba F, Pracucci G, Saia V, Caimano D, Limbucci N, Renieri L, Zini A, Inzitari D, Toni D, et al. Bridging versus direct endovascular therapy in basilar artery occlusion. J Neurol Neurosurg Psychiatry. 2021;92:956–962.

18. Nie X, Wang D, Pu Y, Wei Y, Lu Q, Yan H, Liu X, Zheng L, Liu J, Yang X, et al. Endovascular treatment with or without intravenous alteplase for acute ischaemic stroke due to basilar artery occlusion. Stroke Vasc Neurol. 2022;7:190–199.

19. Kohli GS, Schartz D, Whyte R, Akkipeddi SM, Ellens NR, Bhalla T, Mattingly TK, Bender MT. Endovascular thrombectomy with or without intravenous thrombolysis in acute basilar artery occlusion ischemic stroke: A meta-analysis. J Stroke Cerebrovasc Dis. 2022;31:106847.

20. Ren Y, Churilov L, Mitchell P, Dowling R, Bush S, Yan B. Clot Migration Is Associated With Intravenous Thrombolysis in the Setting of Acute Ischemic Stroke. Stroke. 2018;49:3060–3062.

21. de Havenon A, Alexander MD, Nogueira RG, Haussen DC, Castonguay AC, Linfante I, Johnson MA, Nguyen TN, Mokin M, Zaidat OO. Duration of symptomatic stroke and successful reperfusion with endovascular thrombectomy for anterior circulation large vessel occlusive stroke. J Neurointerv Surg. 2021;13:1128–1131.

